# The negative impact of COVID-19 on working memory revealed using a rapid online quiz

**DOI:** 10.1101/2022.05.20.22275380

**Authors:** Heidi A Baseler, Murat Aksoy, Abayomi Salawu, Angela Green, Aziz UR Asghar

## Abstract

Although coronavirus disease 2019 (COVID-19) affects the respiratory system, it can also have neurological consequences leading to cognitive deficits such as memory problems. The aim of our study was to assess the impact of COVID-19 on working memory function. We developed and implemented an online anonymous survey with a working memory quiz incorporating aspects of gamification to engage participants. 5428 participants successfully completed the survey and memory quiz between 8^th^ December 2020 and 5^th^ July 2021 (68.6% non-COVID-19 and 31.4% COVID-19). Most participants (93.3%) completed the survey and memory quiz relatively rapidly (mean time of 8.84 minutes). Categorical regression was used to assess the contribution of COVID status, age, time post-COVID (number of months elapsed since having had COVID), symptoms, ongoing symptoms and gender, followed by non-parametric statistics. A principal component analysis explored the relationship between subjective ratings and objective memory scores. The objective memory scores were significantly correlated with participants’ own assessment of their cognitive function. The factors significantly affecting memory scores were COVID status, age, time post-COVID and ongoing symptoms. Our main finding was a significant reduction in memory scores in all COVID groups (self-reported, positive-tested and hospitalised) compared to the non-COVID group. Memory scores for all COVID groups combined were significantly reduced compared to the non-COVID group in every age category 25 years and over, but not for the youngest age category (18-24 years old). We found that memory scores gradually increased over a period of 17 months post-COVID-19. However, those with ongoing COVID-19 symptoms continued to show a reduction in memory scores. Our findings demonstrate that COVID-19 negatively impacts working memory function, but only in adults aged 25 years and over. Moreover, our results suggest that working memory deficits with COVID-19 can recover over time, although impairments may persist in those with ongoing symptoms.

## Introduction

Severe acute respiratory syndrome coronavirus (SARS-CoV-2), which causes coronavirus disease 2019 (COVID-19), mainly affects the respiratory system, but can also impact neurological function [1, 2]. Reports in the media have indicated that many people with COVID-19 experience ‘brain fog’ with problems remembering, concentrating, and performing daily tasks. Of growing concern are the number of people suffering continued COVID-19 symptoms for months after infection (‘long COVID’). Scientific investigations have provided evidence that COVID-19 can give rise to cognitive deficits including memory loss [2-6] which can persist for at least six months post-COVID [7]. A systematic review and meta-analysis of individuals with suspected or laboratory-confirmed coronavirus (SARS, MERS, or SARS-CoV-2) has shown that ∼20% had memory impairments in the post-illness phase [8].

Working memory, a form of short-term memory, is the dynamic process by which information is stored and retrieved while performing a task [9-12]. Working memory is critically important in daily living; it is involved in tasks such as problem solving, reasoning, reading comprehension, having a conversation, and is highly correlated with measures of cognitive function [12]. Working memory is short-lasting, has a limited capacity, and declines with age [12-14]. A systematic review of the impact of COVID-19 on cognitive function found thirteen studies which reported deficits in working memory [6].

Researchers investigating the impact of COVID-19 on cognitive abilities have typically utilised researcher-led surveys and tests [5, 15-19]. Using this methodological approach, short-term memory deficits have been reported for COVID-19 in some studies, although these investigations have been limited to small cohorts (6-87 patients) [5, 15, 16, 18, 19]. Despite the usefulness of such survey methods they are time-intensive, require a dedicated investigator, may be subject to selection biases due to issues such as lack of anonymity or ability to travel, limiting the number and breadth of participants recruited. By comparison, online surveys do not require a researcher, can reach a larger and broader sample of the population, and can be completed at the participant’s own convenience and timescale. For example, some online studies of COVID-19 and cognitive function have successfully recruited larger numbers of participants from the general population [20], presenting a range of tasks to test cognition along with other aspects of brain function [20, 21]. However, these online tasks took a relatively long period of time to complete as participants were required to undertake numerous cognitive/memory tests in addition to answering various questionnaires on demographics and other factors. Such long duration online surveys with multiple tasks are likely to increase fatigue levels and decrease completion rates especially in those with COVID-19 or with continued symptoms. In addition, incorporating multiple tasks may have reduced statistical power, led to unknown interactions between tasks, and complicated data interpretation.

The primary aim of our study was to investigate the impact of COVID-19 on working memory function. We designed and utilised an online survey with a single working memory quiz which could be completed rapidly by participants (<15 minutes) via a variety of platforms including smartphones, tablets or PCs. We anonymised the survey and memory quiz to maintain privacy and to increase participant recruitment. To motivate participants, we implemented aspects of gamification into the working memory quiz. In our analysis we evaluate the impact of COVID-19 status (non-COVID, and self-reported, positive tested or hospitalised with COVID), recovery time, COVID-19 symptoms, ongoing symptoms, age and gender on objective memory scores.

## Materials and Methods

### Ethics

The study was carried out in conformity with the Declaration of Helsinki (World Medical Association, 2013)[22]. Local ethical approval was given by the Hull York Medical School Ethics Committee (Reference 20 62). Anyone who was 18 years old and over was invited to complete the survey and memory quiz regardless of whether they had COVID-19. The COVID-19 Online Rapid Objective Neuro-Memory Assessment (CORONA) survey and memory quiz was made publicly available from the 8^th^ of December 2020 and disseminated mainly via direct communication through personal and professional contacts as well as through local intranets and social media. Only participants who gave their active digital informed consent were allowed to complete the survey and memory quiz. The CORONA survey and memory quiz was fully anonymous, no personal contact details were collected, and was in compliance with General Data Protection Regulations (GDPR). The data collected were stored on the Qualtrics platform where all the data were encrypted, and access authorisation was restricted. Participants did not receive any compensation for completing the survey and memory quiz.

### Survey and memory quiz design

The survey/memory quiz was designed to be globally accessible, completed quickly, and engaging in order to maximise recruitment and completion. The online survey and memory quiz was developed using the Qualtrics platform accessed via a University of York licence. The survey was accessible via a Uniform Resource Locator (URL) or a QR code and was designed to work on smartphones, tablets, laptops and personal computers with internet access. Fig 1 gives examples of the survey questions asked and the memory quiz. The survey component consisted of a series of Yes/No questions asking participants about their COVID-19 status (positive test for COVID-19, hospitalisation, self-reported COVID-19 without a positive test), and the month and year when they first had or may have had COVID-19. All participants were then asked to select if they had no symptoms (no symptom group) or if they experienced any of the following symptoms during the COVID-19 pandemic: a high temperature, a new continuous cough, loss of smell or taste, diarrhoea, difficulty breathing, tiredness or other symptoms (symptom group). If any symptoms were selected, participants were then asked which of them they still had (ongoing symptoms group). The next part of the survey asked participants to rate their responses subjectively (using ‘None’, ‘Mild’, ‘Moderate’ or ‘Severe’) to the following 16 questions: ‘*During the COVID-19 pandemic, have you had MORE problems with: Remembering things, Concentrating on simple tasks, Concentrating on complex tasks, Having a conversation, Thinking quickly, Thinking clearly, Getting tired easily after mental effort, Low mood, Fatigue*.’ In addition, we used the same scale to assess whether they had more problems with vision, hearing, smell, taste, touch or pain, balance, and dizziness. Finally, they were asked to select their age range (18-24, 25-34, 35-44, 45-54, 55-64, 65-74, 75-84 and 85+), gender, and country of current residence. No time limit was imposed upon participants to complete the survey questions.

**Fig 1.**
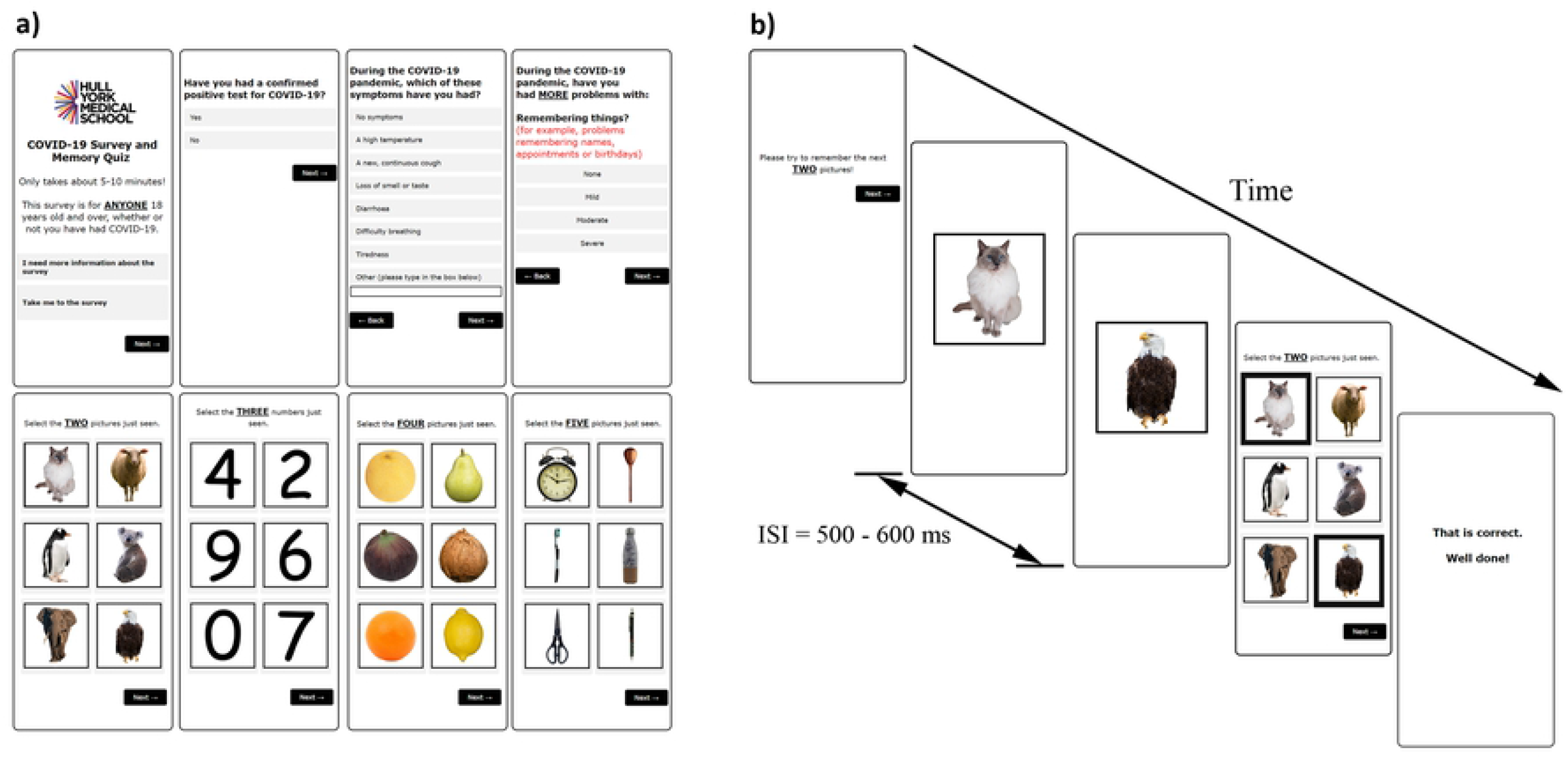
The COVID-19 Online Rapid Objective Neuro-Memory Assessment (CORONA) as presented on a smartphone. a) Selected sample questions from the survey, and image examples of the animal, number, fruit and object categories presented in the memory quiz. b) Example showing the timing of a single trial presentation during the objective memory quiz.

We developed a visual working memory/recall task based upon modifications of the established and widely utilised visual simple picture span and the digit span tasks [23, 24]. The working memory quiz required minimal levels of English language proficiency and education level to complete. The images and categories we selected were easily identifiable and non-threatening. We applied elements of gamification into our working memory quiz including a simple to understand task, familiar and engaging images, increasing levels of difficulty, feedback on answers, and reward via points scored. Four categories of images comprising animals, numbers, fruits, and objects were presented to participants (Fig 1a). The stimuli set comprised of 55 unique images, consisting of animals (15 unique images), fruits (15), objects (15) and single-digit whole numbers (10; 0 to 9). Colour photographs for three of the categories (animals, fruits and objects) were taken from two open-source image databases (Pixabay, https://pixabay.com/ and Unsplash, https://unsplash.com/) and from the researcher’s personal image gallery. All images were edited using Adobe Photoshop CC 2015 software. Images of single-digit whole numbers were generated by the researchers using Comic Sans MS regular font. The backgrounds of all images were removed, and each image was rescaled and centralised onto a 500 × 500 pixel white background square with a black outline.

After completing the survey questions, participants were next asked to complete the objective working memory quiz. To improve concentration and compliance, participants were instructed to ‘*Please complete the quiz in a QUIET place WITHOUT DISTRACTIONS*.’ The animal category was presented first to encourage participants to engage with the memory task, followed by either the number or fruit categories (randomised). Based upon pilot testing, the object category was found to be relatively more difficult, so we opted to present this category last. For each image category, two different random images were shown consecutively. Each image was presented for 500ms and the interstimulus interval was randomly varied between 500-600ms (Fig 1b). Next, a grid of six images was presented and participants were instructed to select the two images viewed previously. No time limit was imposed upon participants to make a response. To keep participants engaged and challenged, this process was repeated with participants tasked to recall three, four and five images shown resulting in a total of four trials per category. A score of 1 was given for each correct trial, yielding a possible maximum objective memory score of 16. Participants were given immediate feedback after each trial, either ‘*That is correct. Well done!*’ or ‘*That is not correct*.’ Following completion of the memory quiz, participants were given their total percentage correct score along with the percentage scores for each image category.

### Data analysis

Statistical analysis was performed using SPSS (Version 27.0, IBM Corporation, Armonk, N.Y., USA). To determine the relative contribution of each independent variable to the objective memory scores we used categorical regression (Optimal scaling-CATREG, SPSS). First, we assessed the contributions of COVID status (x2 groups: non-COVID, COVID), symptoms (2 groups: symptoms, no symptoms), ongoing symptoms (2 groups: ongoing symptoms, no ongoing symptoms), age (8 groups: 18-24, 25-34, 35-44, 45-54, 55-64, 65-74, 75-84 and 85+) and gender (2 groups: female, male). A second categorical regression was performed on the COVID group only to evaluate the effects of time post-COVID (17 groups: ≤1 to 17 months) along with symptoms, ongoing symptoms, age and gender. Based upon the categorical regression analysis, we next examined the impact of the significant individual variables on objective memory scores utilising non-parametric statistics: Mann Whitney U test and Kruskal-Wallis test with Dunn’s pairwise comparisons (Bonferroni corrected). Finally, we explored the relationship between subjective ratings in the survey and objective memory quiz scores. To reduce dimensionality of the 16 subjective questions, a principal component analysis (PCA) with orthogonal (varimax) rotation was performed. Relationships between variable pairs were evaluated using the Kendall’s tau-b correlation coefficients. All testing was two-tailed, and *p* values less than 0.05 were considered to be statistically significant.

## Results

### Participant demographics

A total of 5428 participants fully completed the CORONA survey and memory quiz between 8^th^ December 2020 and 5^th^ July 2021. Of these, 3722 participants reported not having had COVID-19, and 1706 participants reported having had COVID-19 between January 2020 and July 2021. Table 1 lists the age, gender and country of residence of the participants recruited. Participants were recruited in each age range, with the highest number recruited in the 45-54 age group (1385 participants). Most of the study participants selected the United Kingdom (UK) as their current place of residence (4957 participants), and 471 participants were recruited outside the UK from 43 different countries. The duration to complete the entire survey and memory quiz was recorded in 5391 participants (37 participants did not click on the closing screen which meant that the duration time was not recorded). Of these, 5028 participants (93.3%) completed the survey and memory quiz within 15 minutes (Mean ± SEM, 8.84 ± 0.03 minutes; range 5.05-15 minutes).

**Table 1.** Participant demographics.

| **Age (Years)** | **N** |
| --- | --- |
| 18 - 24 | 657 |
| 25 - 34 | 687 |
| 35 - 44 | 978 |
| 45 - 54 | 1385 |
| 55 - 64 | 1149 |
| 65 - 74 | 458 |
| 75 - 84 | 109 |
| 85+ | 5 |
| **Gender** |  |
| Female | 4313 |
| Male | 1088 |
| Other | 11 |
| Prefer not to say | 16 |
| **Countries** |  |
| UK | 4957 |
| Non-UK | 471 |

### Categorical regression and objective memory scores

For all participants (non-COVID and COVID groups), a categorical regression was used to determine the contributions of COVID status, COVID symptoms, ongoing symptoms, age and gender on memory scores. The analysis generated an overall regression coefficient of multiple determination of R^2^ = 0.035, and p < 0.001. Table 2 shows that there was a significant effect for COVID status, ongoing symptoms (while completing the survey and memory quiz) and age (all values of p < 0.001), but not for symptoms (anytime during the pandemic) and gender (p > 0.05). The standardised regression coefficient for each independent variable showed that the memory quiz scores are mostly affected by age (51.1%), followed by COVID status (26.9%), and ongoing symptoms (24.1%).

**Table 2.**
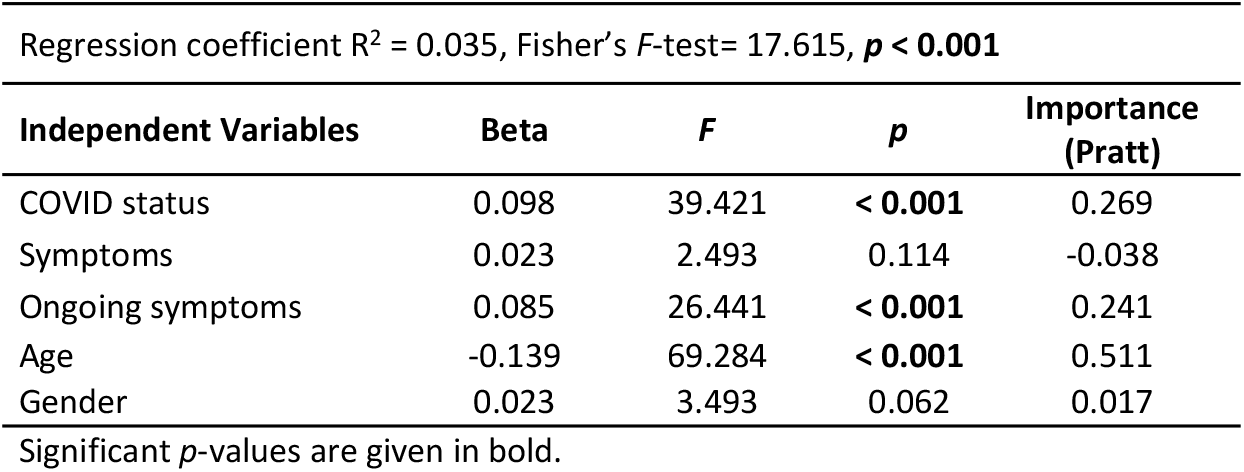
Categorical regression analysis and objective memory scores for non-COVID and COVID participants combined.

| Regression coefficient $R^2 = 0.035$, Fisher's $F$-test= 17.615, $p < 0.001$ | | | | |
| --- | --- | --- | --- | --- |
| Independent Variables | Beta | $F$ | $p$ | Importance (Pratt) |
| COVID status | 0.098 | 39.421 | **< 0.001** | 0.269 |
| Symptoms | 0.023 | 2.493 | 0.114 | -0.038 |
| Ongoing symptoms | 0.085 | 26.441 | **< 0.001** | 0.241 |
| Age | -0.139 | 69.284 | **< 0.001** | 0.511 |
| Gender | 0.023 | 3.493 | 0.062 | 0.017 |

Significant $p$ -values are given in bold.

For the COVID group only, a categorical regression was used to determine the contributions of time post-COVID (recovery time), COVID-19 symptoms, ongoing symptoms, age and gender on memory scores. The regression coefficient of multiple determination was R^2^ = 0.04, with p < 0.001. Table 3 shows that there was a significant effect of time post-COVID, ongoing symptoms, and age (all values of p < 0.05), but not for symptoms and gender (p > 0.05). The standardised regression coefficient for each independent variable showed that the memory quiz scores are mostly affected by age (45.5%), followed by ongoing symptoms (34.2%), and time post-COVID (13.8%).

**Table 3.** Categorical regression analysis and objective memory scores for participants in the COVID group only.

| Regression coefficient $R^2 = 0.04$, Fisher's $F$-test= 5.180, $p < 0.001$ | | | | |
| --- | --- | --- | --- | --- |
| Independent Variables | Beta | $F$ | $p$ | Importance (Pratt) |
| Time post-COVID | 0.075 | 3.972 | **0.003** | 0.138 |
| Symptoms | 0.026 | 3.341 | 0.068 | 0.039 |
| Ongoing symptoms | 0.098 | 14.792 | **< 0.001** | 0.342 |
| Age | -0.132 | 35.298 | **< 0.001** | 0.455 |
| Gender | 0.026 | 2.189 | 0.139 | 0.026 |

Significant $p$ -values are given in bold.

Since the categorical regressions revealed that the main factors affecting objective memory scores were COVID status, age, time post-COVID and ongoing symptoms, we evaluate these factors in more detail below. We used non-parametric statistics in all these subsequent analyses as a Kolmogorov-Smirnov test revealed that objective memory scores were not normally distributed [D(5428) = 0.25, p < 0.05].

### Effect of COVID status on objective memory scores

Fig 2a shows that the objective memory scores are significantly reduced in the COVID group [Mean ± SEM (M) = 14.74 ± 0.04] compared to the non-COVID group (M = 15.00 ± 0.02, Mann – Whitney U = 2902729.50, z = –5.36, p < 0.001, r = –0.07). Next, we divided the COVID group into sub-groups: those who were not tested but suspected they may have had COVID (self-reported), those who tested positive for COVID, and those who were hospitalised with COVID. Fig 2b shows that objective memory scores were highest in the non-COVID group, but significantly reduced in each of the COVID sub-groups, with the lowest scores in the hospitalised COVID group. A Kruskal-Wallis test revealed a significant difference between the mean memory scores across groups [H(3) = 43.42, p < 0.001]. Dunn’s pairwise comparison tests with Bonferroni correction revealed significant differences between the non-COVID group (M = 15.00 ± 0.02) and the self-reported (M = 14.80 ± 0.05, p = 0.002), positive-tested (M = 14.73 ± 0.06, p < 0.001), and hospitalised (M = 13.46 ± 0.41, p < 0.001) COVID groups. In addition, there was significant differences between the self-reported (M = 14.80 ± 0.05) and hospitalised (M = 13.46 ± 0.41, p = 0.001) COVID groups, and between positive-tested (M = 14.73 ± 0.06) and hospitalised (M = 13.46 ± 0.41, p = 0.002) COVID groups. There was no significant difference between positive-tested (M = 14.73 ± 0.06) and self-reported (M = 14.80 ± 0.05, p = 1.000) COVID groups.

**Fig 2.**
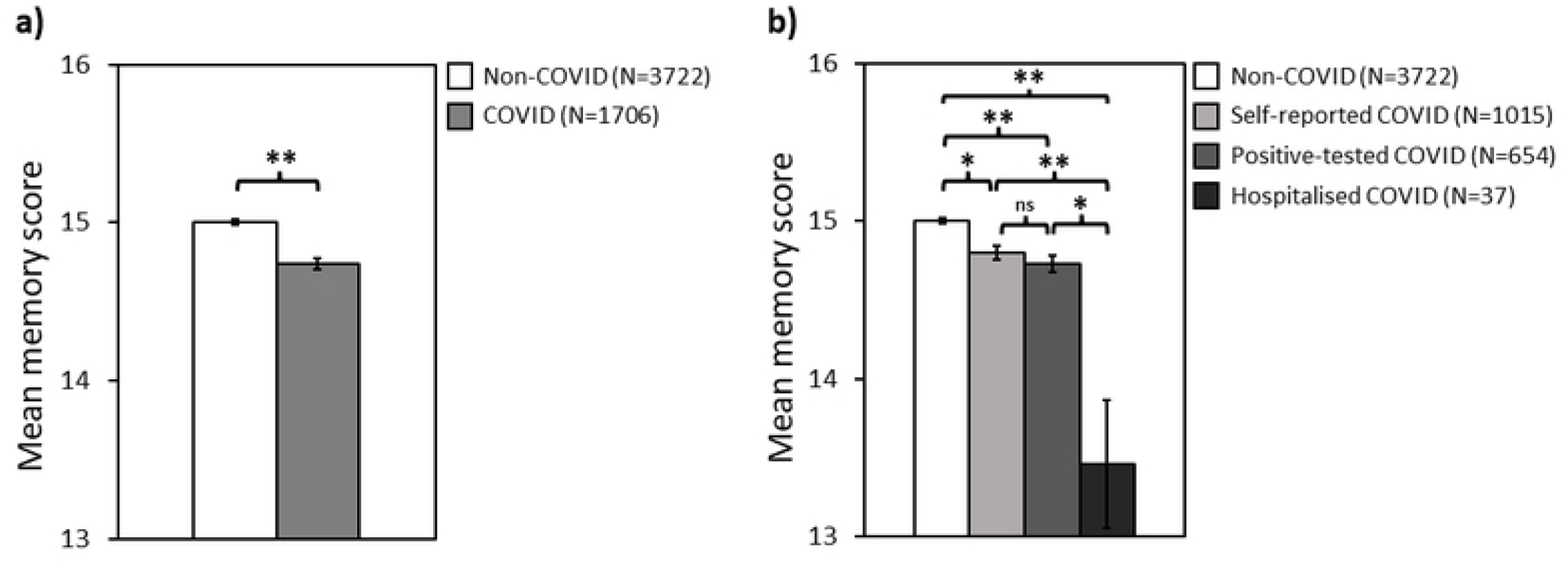
The effect of COVID status on objective memory scores. **a)** Non-COVID group *versus* all COVID groups combined, and **b)** Non-COVID group compared to each COVID sub-group. All differences between group pairs are significant except between self-reported COVID and positive-tested COVID. Error bars represent standard error of the mean. **\*** *p < 0*.*01*, **\*\*** *p < 0*.*001*, **ns:** no significant difference.

### Effect of age on objective memory scores

We found that objective memory scores decreased with age for both the non-COVID and COVID groups. A Kendall’s tau-b correlation showed a negative relationship between mean memory score and age for both the non-COVID (τb = –0.118, p < 0.001) and COVID (τb = – 0.114, p < 0.001) groups across all eight age categories. All participants above 55 years old were combined to increase sample size due to the lower numbers of participants who completed the memory quiz in the higher age categories. Fig 3 shows the effect of age on memory scores for COVID and non-COVID groups for five age categories. A Kruskal-Wallis test revealed that the mean memory score differed significantly across age groups for non-COVID and COVID groups combined [H(4) = 88.77, p < 0.001]. Planned comparisons with Bonferroni correction revealed that the mean memory score of the COVID group was significantly lower compared to the non-COVID group in each age group 25 years old and over (Mann – Whitney U, all adjusted values of p < 0.05, Table 4). However, within the age range 18-24, there was no significant difference between the mean memory scores for COVID and non-COVID groups (Mann – Whitney U, p > 0.05).

**Table 4.** Mann-Whitney U comparisons for objective memory scores between COVID and non-COVID groups for each age range.

| Age Groups | N | Mean Rank | Z | U | p | Adjusted p* |
| --- | --- | --- | --- | --- | --- | --- |
| 18-24 |  |  |  |  |  |  |
| COVID | 259 | 329.80 | -0.095 | 51333.5 | 0.924 | >1.000 |
| Non-COVID | 398 | 328.48 |  |  |  |  |
| 25-34 |  |  |  |  |  |  |
| COVID | 244 | 311.21 | 3.449 | 62046.0 | 0.001 | 0.006* |
| Non-COVID | 442 | 362.06 |  |  |  |  |
| 35-44 |  |  |  |  |  |  |
| COVID | 348 | 443.66 | 3.993 | 125571.0 | <0.001 | <0.001* |
| Non-COVID | 630 | 514.82 |  |  |  |  |
| 45-54 |  |  |  |  |  |  |
| COVID | 433 | 636.93 | 3.709 | 230387.5 | <0.001 | <0.001* |
| Non-COVID | 952 | 718.50 |  |  |  |  |
| 55+ |  |  |  |  |  |  |
| COVID | 422 | 801.79 | 2.943 | 299077.5 | 0.003 | 0.018* |
| Non-COVID | 1299 | 880.24 |  |  |  |  |

\* Represents significant difference between COVID and non-COVID groups.

**Fig 3.**
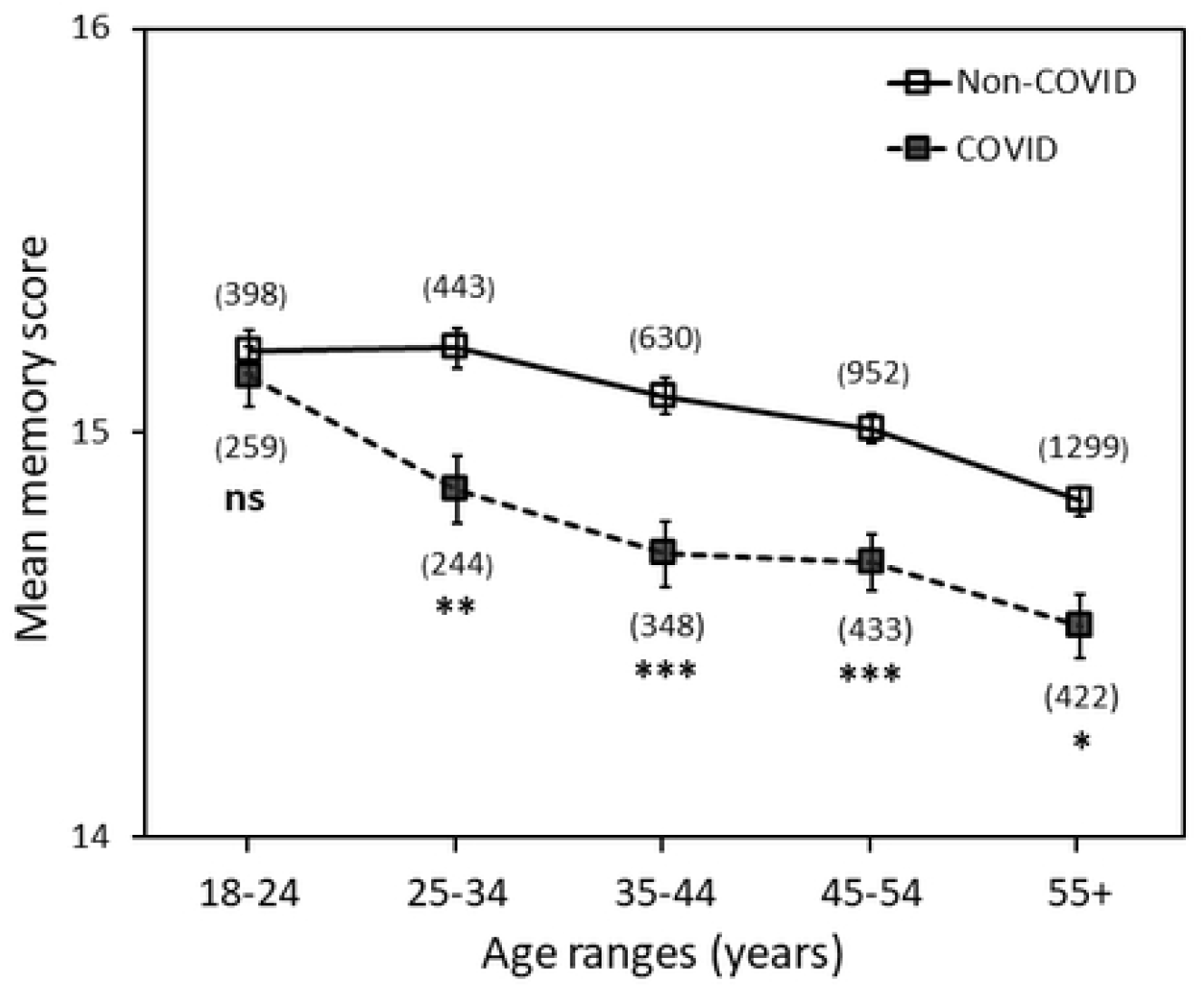
The effect of age on objective memory scores in the COVID and non-COVID groups. Sample sizes are given in brackets for each data point. Error bars represent the standard error of the mean. Asterisks indicate significant differences compared to the non-COVID group for each age category. **\*** *p < 0*.*05*, **\*\*** *p < 0*.*01*, ***\*\*\**** *p < 0*.*001*, **ns**, no significant difference.

### Effect of time post-COVID on objective memory scores

We found that within the COVID group, objective memory scores increased with time (number of months) post-COVID. A Kendall’s tau-b correlation revealed a positive correlation between the number of months (≤1 to 17) since having had COVID and objective memory scores (τb = 0.04, p = 0.037). Fig 4 illustrates the change in objective memory scores since having had COVID. Table 5 lists the results of planned individual comparisons for the objective memory scores between each post-COVID group and the non-COVID group. Mann-Whitney U tests revealed that there were significant differences between the non-COVID group and the 0-3 month post-COVID group, and between the non-COVID group and the 8-11 month post-COVID group when correcting for multiple comparisons (Table 5). Scores for the group 4-7 months post-COVID were lower than those in the non-COVID group, but this was not significant after correcting for multiple comparisons. There was no significant difference in memory scores for the 12+ month group compared to the non-COVID group, whether or not correcting for multiple comparisons (Table 5).

**Table 5.** Mann-Whitney U comparisons for objective memory scores between the non-COVID group for each time period post-COVID.

| Groups | N | Mean Rank | Z | U | p | Adjusted p* |
| --- | --- | --- | --- | --- | --- | --- |
| **Non-COVID**<br>**compared to:** | 3722 | 1991.11 |  |  |  |  |
| **0-3 months post-COVID** | 501 | 1909.79 | -4.184 | 831053.50 | < 0.001 | **< 0.001\*** |
| **4-7 months post-COVID** | 204 | 1809.03 | -2.119 | 348132.00 | 0.034 | 0.204 |
| **8-11 months post-COVID** | 754 | 2083.89 | -3.812 | 1286614.50 | < 0.001 | **< 0.001\*** |
| **12+ months post-COVID** | 247 | 1892.95 | -1.382 | 436929.50 | 0.167 | 1.002 |

\* Represents significant difference between the post-COVID and non-COVID group.

**Fig 4.**
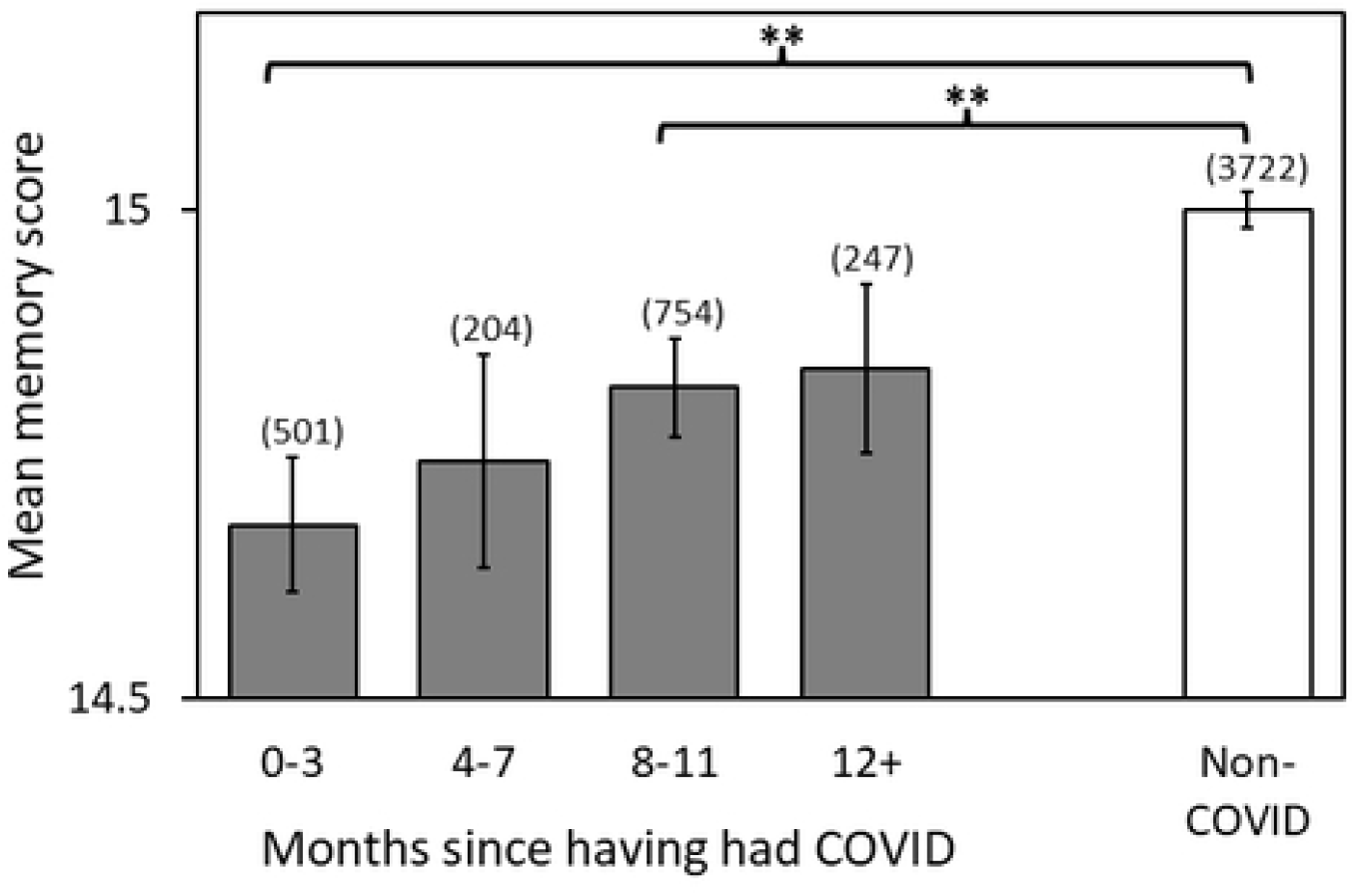
Objective memory scores over time post-COVID. Sample sizes are given in brackets for each data point. Error bars represent the standard error of the mean. Asterisks indicate significant differences compared to the non-COVID group, **\*\*** *p < 0*.*001* (corrected for multiple comparisons). Note that for the 12+ group only, there was no significant difference compared to the non-COVID group whether or not adjusted for multiple comparisons (see Table 5).

### Effect of ongoing symptoms on objective memory scores

Fig 5a shows that, irrespective of whether participants had COVID or not, there was a significant reduction in the mean memory scores for those with ongoing symptoms compared to those without ongoing symptoms (no ongoing symptoms: M = 14.99 ± 0.03; ongoing symptoms: M = 14.72 ± 0.04; U = 1164131.50, z = 3.75, p < 0.001, r = 0.07). Fig 5b shows the participants with and without ongoing symptoms subdivided into the non-COVID and COVID groups. Of the COVID group, 797 participants had no ongoing symptoms, and 811 participants (∼50%) reported having ongoing symptoms while completing the survey and memory quiz: high temperature, new continuous cough, loss of smell or taste, diarrhoea, difficulty breathing, tiredness or other self-reported symptoms. A Kruskal-Wallis test found a significant effect on the mean memory score [H(3) = 43.36, p < 0.001]. Dunn’s pairwise comparisons with Bonferroni correction revealed that the mean memory scores of the COVID group with ongoing symptoms (M = 14.53 ± 0.06) were significantly lower compared to non-COVID with ongoing symptoms (M = 14.93 ± 0.05, p < 0.001) or without symptoms (M = 15.09 ± 0.04, p < 0.001), and compared to the COVID group with no ongoing symptoms (M = 14.92 ± 0.04, p < 0.001). There were no statistically significant differences found for the other comparisons (all values of p > 0.05).

**Fig 5.**
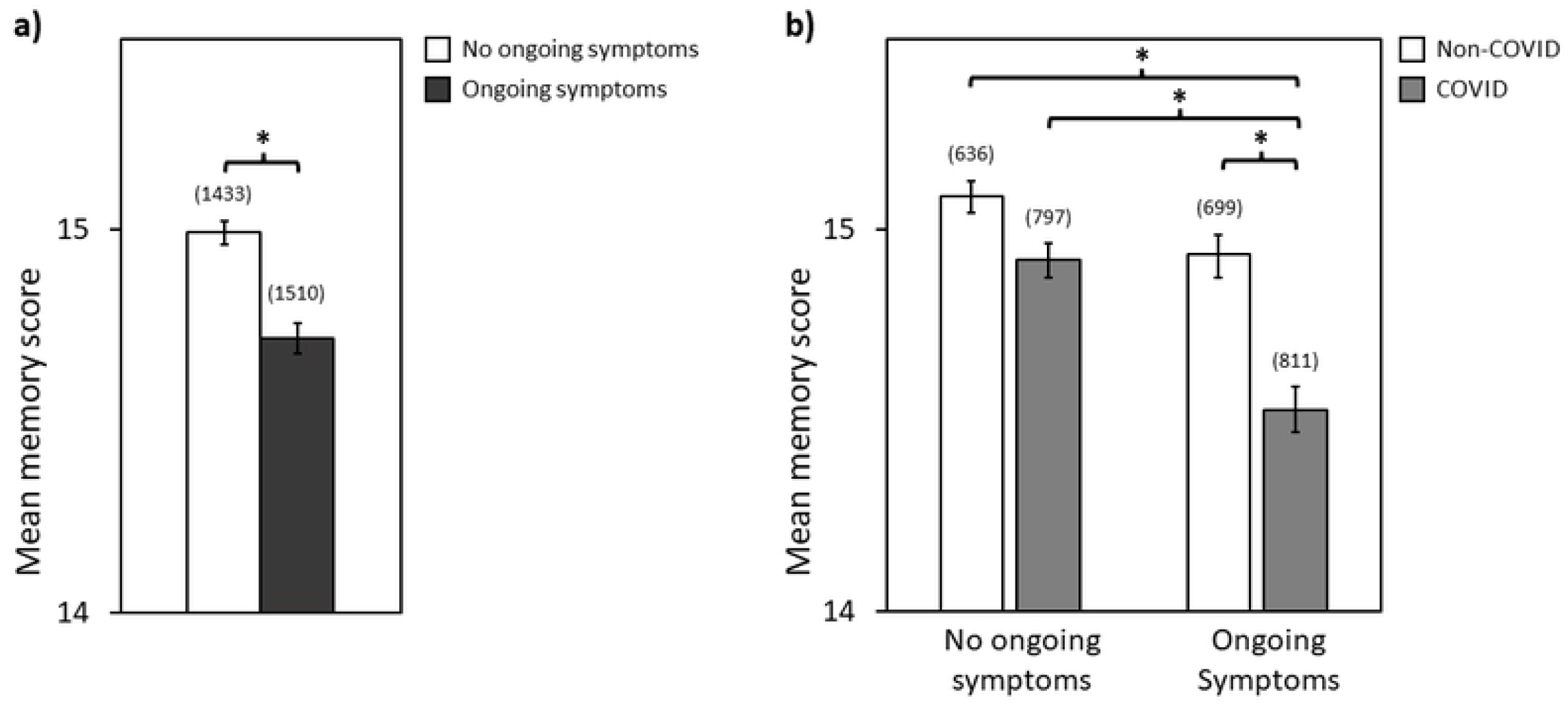
The effect of ongoing symptoms on objective memory scores. **a)** All participants (non-COVID and COVID combined), with and without ongoing symptoms, and **b)** non-COVID and COVID participants with and without ongoing symptoms (high temperature, new continuous cough, loss of smell or taste, diarrhoea, difficulty breathing, tiredness or other self-reported symptoms). Error bars represent the standard error of the mean. **\*** *p < 0*.*001*.

### Correlations between subjective ratings and objective memory scores

To reduce dimensionality and assess the intercorrelation between the subjective ratings of the 16 survey questions, a principal component analysis (PCA) with orthogonal (varimax) rotation was performed on. The Kaiser-Meyer-Olkin measure verified the sampling adequacy for the analysis, KMO = 0.913, and all KMO values for individual items were > 0.670, which is well above the acceptable limit of .5 [25]. Bartlett’s test of sphericity, χ^2^ (120) = 50790.90, *p* < 0.001, indicated that correlations between items were sufficiently large for PCA. An initial analysis was run to obtain eigenvalues for each component in the data. Three components had eigenvalues over Kaiser’s criterion of 1 and in combination explained 63.49% of the variance. Table 6 shows the factor loadings after rotation, sorted in order from highest to lowest loadings for each component. The questions that cluster on the same components suggest that component 1 primarily represents cognitive function, component 2 primarily reflects sensory function other than smell and taste, and component 3 primarily captures smell and taste function. Next, we determined the relationship between participants’ subjective ratings for each of these three components with objective memory quiz scores. For all participants (non-COVID and all COVID groups combined), a Kendall’s tau-b correlation showed a negative relationship between the objective mean memory score and the mean subjective rating for each component (cognitive component, τb = –0.052 p < 0.001; sensory component, τb = –0.086 p < 0.001; smell/taste component, τb = –0.067 p < 0.001). In other words, participants who reported more severe subjective ratings had lower objective memory scores. For each principal component (cognitive, sensory, and smell/taste), Fig 6 illustrates the average subjective ratings plotted against the objective memory scores for the non-COVID, self-reported COVID, positive-test COVID and hospitalised COVID groups. The figure demonstrates that the non-COVID group reported the least severe ratings, while those who were hospitalised with COVID reported the most severe ratings.

**Table 6.** Principal component analysis of subjective questions in the CORONA survey (N=5428).

| Subjective Questions<sup>a</sup> | Principal Components<sup>b</sup> |  |  |
| --- | --- | --- | --- |
|  | Cognitive | Sensory | Smell/ Taste |
| Thinking clearly | **0.835** | 0.238 | 0.087 |
| Concentrating on complex tasks | **0.832** | 0.193 | 0.082 |
| Thinking quickly | **0.799** | 0.256 | 0.094 |
| Concentrating on simple tasks | **0.790** | 0.174 | 0.081 |
| Having a conversation | **0.762** | 0.264 | 0.067 |
| Getting tired easily after mental effort | **0.736** | 0.271 | 0.115 |
| Remembering things | **0.716** | 0.279 | 0.070 |
| Fatigue | **0.706** | 0.262 | 0.149 |
| Low mood | **0.646** | 0.129 | 0.029 |
| Balance | 0.222 | **0.757** | 0.161 |
| Dizziness | 0.231 | **0.702** | 0.222 |
| Vision | 0.242 | **0.631** | -0.012 |
| Hearing | 0.175 | **0.605** | -0.029 |
| Touch or pain | 0.248 | **0.598** | 0.219 |
| Taste | 0.123 | 0.153 | **0.936** |
| Smell | 0.126 | 0.144 | **0.933** |
| Eigenvalues | 7.127 | 1.801 | 1.231 |
| % of variance | 44.544 | 11.253 | 7.695 |

<sup>a</sup>Each subjective question was preceded by the introductory phrase “During the COVID-19 pandemic, have you had ***MORE*** problems with:”
<sup>b</sup>Rotated factor loadings over 0.50 appear in bold.

**Fig 6.**
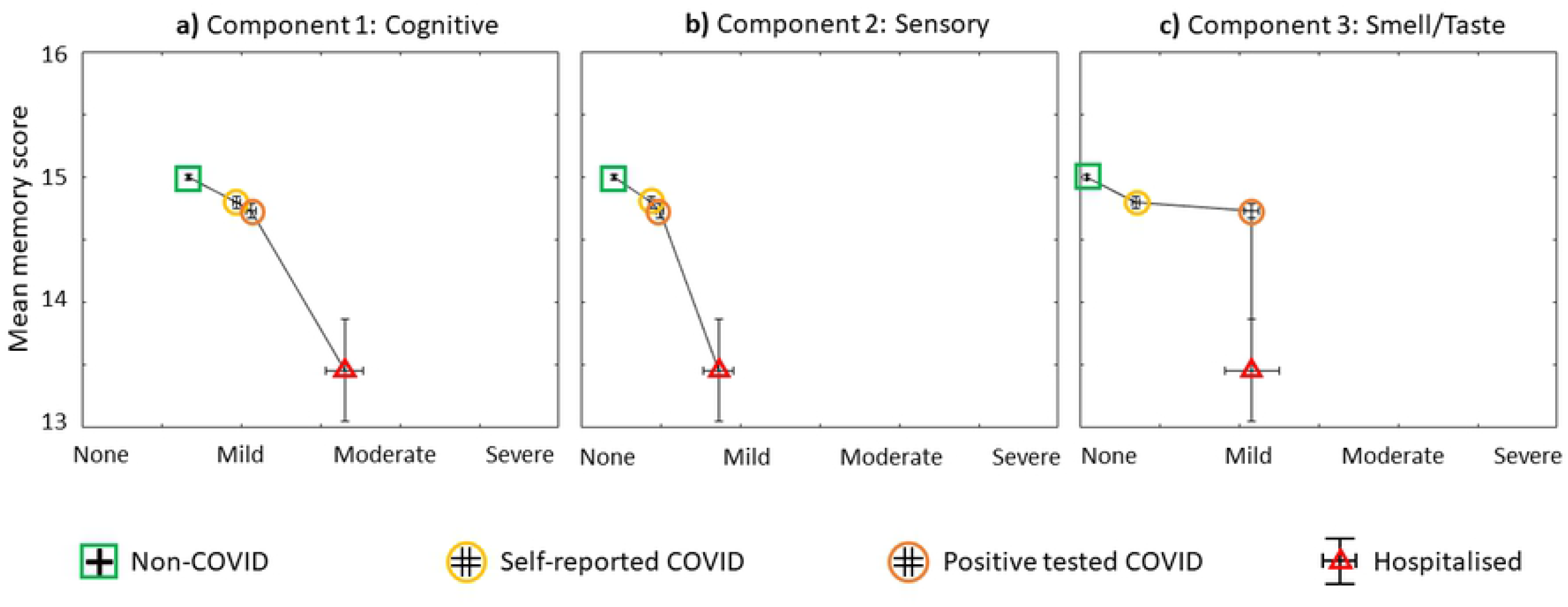
Subjective ratings *versus* objective memory scores for each principal component. **a)** Cognitive, **b)** Sensory and **c)** Smell/Taste components. The mean objective memory score is plotted against the mean subjective rating for each of the following groups: non-COVID (N=3722), self-reported COVID (N=1015), positive-tested COVID (N=654) and hospitalized COVID (N=37). Vertical error bars represent the standard error of the mean for memory scores, horizontal error bars represent the standard error of the mean for subjective ratings across each group.

## Discussion

Our online approach led to the successful recruitment of 5428 participants over a period of seven months (68.6% non-COVID-19 and 31.4% with COVID-19). Our study indicates that the factors most affecting objective memory scores were COVID status, age, time post-COVID and ongoing symptoms. The main finding was that objective memory scores were significantly reduced in the self-reported, positive-tested and hospitalised COVID groups compared to the non-COVID group. These results suggest that COVID-19 affects working memory function. With respect to age, we found that memory scores for the COVID group were reduced compared to the non-COVID group in every age category 25 years and over, but not for the youngest age category (18-24 years old). For the COVID group, memory scores increased as the number of months elapsed since having had COVID-19, indicating that working memory deficits can recover over time. However, scores were reduced for those with ongoing COVID-19 symptoms suggesting that working memory impairments can persist in those who continue to suffer the effects of COVID-19.

Our key discovery was a significant reduction in memory quiz scores in the COVID group (N=1706) compared to the non-COVID group (N=3722). This finding provides quantitative evidence that COVID-19 negatively impacts working memory function. The reduction in memory scores is consistent with the short-term memory deficits reported in smaller scale studies using screening questionnaires, paper-based assessments or in-person testing [5-7, 15, 16, 18, 19, 26]. Given the importance of working/short-term memory for everyday tasks, such as having a conversation, decision-making, reading comprehension, performing a sequence of actions, it is reasonable to conclude that COVID-19-induced working memory deficits may compromise performance in daily life.

Although the difference in objective memory scores between the COVID and non-COVID group was statistically significant it was small. However, a small change in working memory might substantially impact performance of everyday tasks. To support this, we found that objective memory scores correlated strongly with participants’ own assessment of their cognitive function; participants who reported more severe subjective ratings for cognitive questions had lower objective memory scores. Moreover, participants who were hospitalised for COVID-19 showed a marked drop in objective scores and an increase in severity in the subjective ratings. In line with these results, one study reported that 55% of hospitalised patients had working memory deficits [18], and others found that working memory impairments can persist 2-4 months after hospital discharge [19, 27]. Moreover, other investigations of hospitalised patients found a greater deficit in cognitive function with increasing COVID severity [5, 8, 15, 16].

In contrast to our findings, a large-scale study found that complex cognitive functions (reasoning, planning and problem-solving tasks) were generally more affected while simpler cognitive functions (basic working memory tasks) were relatively spared in those with COVID-19 [20]. Nevertheless, this study did find that performance on a spatial span working memory task was significantly affected in the least severe COVID group (symptoms without respiratory symptoms). In another study with more limited participant numbers, short-term memory was compared between the COVID (N=36) and control (N=44) groups, and found no significant difference in accuracy for either word or object immediate memory tasks [21]. However, these studies required participants to complete a number of different cognitive tasks, potentially increasing the chance of false negative results. In addition, multiple cognitive tests may have unintended interactions thereby affecting data interpretation. Performing a battery of tests could also lead to participant fatigue and boredom. To avoid such unintended interactions, in the current study we used a single dedicated working memory task, making interpretation straightforward and uncontaminated by other tasks.

In our study we found that objective memory scores declined with age regardless of COVID status. This shows that our memory quiz is sensitive to the well documented steady decline in working memory beyond early adulthood as has been reported in a range of other memory tasks [13, 14]. Interestingly, the memory scores for the COVID group were significantly reduced compared to the non-COVID group but only in participants 25 years old and over. There was no significant difference in the memory scores between the COVID and non-COVID groups for those in the 18-24 age group. These results reveal that COVID-19 reduces working memory in most adults except for those in the youngest age group (18-24 years old). While previous studies have reported that cognitive function is impaired in COVID-19 participants regardless of age, either controlling for age or using age-matched samples [16, 20, 21], our study shows that it is important to consider age. We recommend that future investigations of COVID-19 on working memory should not simply factor out age but examine the relationship between COVID-19 and individual ages. For example, studies could determine which factors protect younger adults from memory deficits with COVID-19, with the aim of designing interventions to protect older adults.

In terms of recovery from COVID-19, objective memory scores in the COVID group increased in the months following COVID. This suggests that working memory decline due to COVID-19 is not permanent but does have the capacity to improve over time. In addition, our data indicates that it takes approximately 12 months to recover from the decline in working memory to the point that scores are not significantly different to those who do not have COVID-19. Similarly, a steady recovery over 12 months was found in a study of COVID-19 survivors (N=24) using a delayed episodic memory task [21].

Approximately 50% of our COVID group reported having ongoing COVID symptoms while completing the survey and memory quiz. This substantial percentage has implications on the burden upon people with COVID-19 symptoms, health service providers and society. Similar ongoing symptoms have been reported in those with long COVID [26, 28]. For the COVID group only, we found that ongoing symptoms resulted in a significant decrease in memory scores compared to participants without ongoing symptoms. This result demonstrates that working memory is affected in those with COVID-19 and with ongoing symptoms. In support of our finding, a questionnaire study reported a ∼50% probability of persistent memory issues with COVID-19 [7]. Moreover, in another study of long COVID, patients with persistent symptoms performed significantly worse on a working memory task compared to a normative group [26].

One limitation with our survey and memory quiz was that it was not conducted under controlled laboratory conditions. For instance, the presence of a researcher may encourage a participant to take the survey more seriously and answer the questions accurately. However, requiring the presence of a researcher would make the study more practically challenging and would limit the number of participants that could be recruited. Furthermore, the presence of a researcher does not necessarily ensure that participants will complete all aspects of the survey and memory quiz accurately. It is also possible that participants may have been distracted during our memory quiz thereby affecting the scores. We attempted to mitigate against this possibility by instructing participants to complete the quiz in a quiet place without distractions. Nevertheless, the large number of participants in both the COVID and non-COVID groups was able to provide sufficient power to reveal a highly significant difference.

In our study, 31.4% of participants reported having had COVID-19 between January 2020 and July 2021. These included those who self-reported, tested positive and were hospitalised with COVID-19. Since COVID-19 tests were not commonly available in the earlier stages of the pandemic, we allowed participants to self-report whether they thought they may have had COVID-19. We cannot be certain whether these participants in fact had COVID-19. However, we found no significant difference in memory scores between the self-reported COVID group and those who tested positive for COVID, whereas there was a significant difference in memory scores between the self-reported COVID group and the non-COVID group. Therefore, it would be reasonable to consider that most self-reports of COVID-19 are likely to be correct.

The majority of participants (93.3%) were able to finish our survey and the memory quiz within a relatively short period (mean time of 8.84 minutes). In contrast, most other investigations of COVID-19 presented multiple cognitive tasks and questionnaires [6, 7, 18-21]. For instance, participants were required to complete nine [20] or twelve [21] cognitive tasks in addition to multiple questionnaires and demographic questions. In another study, participants were required to answer 257 questions taking a median time of ∼70 minutes to complete [7]. However, using a vigilance task to monitor attention levels, it has been shown that accuracy decreases and fatigue increases over a period of ∼9 minutes particularly in those who have had COVID-19 [21]. Since our survey and single memory task takes a relatively short time to complete, it has the potential for utility in a broader range of the population. For example, it could be used to test people with limited attention spans or those with physical or cognitive deficits who might find long testing periods fatiguing and challenging including those with long COVID.

While our study provides evidence that working memory is negatively impacted by COVID-19, the underlying mechanisms for this are unknown. Several pathophysiological mechanisms underlying the neurological impact of COVID-19 have been proposed, including neuroinflammation, vascular dysfunction, coagulopathy, and pre-existing co-morbidities [1, 3, 29, 30]. It is possible that some of these mechanisms may affect brain areas involved in working memory. Structural and functional alterations in the brain could be investigated using techniques such as structural/functional MRI, electroencephalography, magnetoencephalography, and functional near-infrared spectroscopy. A recent MRI study comparing longitudinal brain images between COVID-19 patients and controls found structural changes in several brain areas primarily involved in olfactory function [31]. However, the links between pathophysiological mechanisms, brain changes, and deficits in working memory function remain to be elucidated.

## Data Availability

The data for the study will be made available in the Supporting Information after manuscript acceptance.

## Acknowledgments

We thank the various Hull York Medical School students for their help in the survey and memory quiz dissemination, and the initial data analysis as part of their Scholarship and Special Interest Programme including: Seren Adams, Jonathon Austin, Lauren Farr, Aya Hammad, Zack Hudson, Aminah Hussain, Tayabah Iftikhar, Nusrat Islam, Jamie McAllister, Thomas Panter, Shayan Saadat, Catherine Woods, Urooj Zahid.

## Author Contributions

**Conceptualization:** Aziz UR Asghar, Heidi A Baseler, Abayomi Salawu

**Data curation:** Aziz UR Asghar, Murat Aksoy

**Formal analysis:** Murat Aksoy, Heidi A Baseler, Aziz UR Asghar

**Investigation:** Aziz UR Asghar, Heidi A Baseler, Murat Aksoy

**Project Administration:** Aziz UR Asghar, Heidi A Baseler, Murat Aksoy

**Methodology:** Aziz UR Asghar, Murat Aksoy, Heidi A Baseler, Abayomi Salawu, Angela Green

**Resources:** Aziz UR Asghar, Murat Aksoy, Heidi A Baseler

**Supervision:** Aziz UR Asghar, Heidi A Baseler, Murat Aksoy, Abayomi Salawu, Angela Green

**Visualization:** Heidi A Baseler, Murat Aksoy, Aziz UR Asghar

**Writing – original draft:** Aziz UR Asghar, Heidi A Baseler

**Writing – review & editing:** Aziz UR Asghar, Heidi A Baseler, Murat Aksoy, Abayomi Salawu, Angela Green

## References

1. Koralnik IJ, Tyler KL. COVID-19: A Global Threat to the Nervous System. Ann Neurol. 2020;88(1):1–11. Epub 2020/06/09. doi: 10.1002/ana.25807. PubMed PMID: 32506549.

2. Fotuhi M, Mian A, Meysami S, Raji CA. Neurobiology of COVID-19. J Alzheimers Dis. 2020;76(1):3–19. Epub 2020/06/17. doi: 10.3233/JAD-200581. PubMed PMID: 32538857.

3. Spudich S, Nath A. Nervous system consequences of COVID-19. Science. 2022;375(6578):267–9. Epub 2022/01/21. doi: 10.1126/science.abm2052. PubMed PMID: 35050660.

4. Daroische R, Hemminghyth MS, Eilertsen TH, Breitve MH, Chwiszczuk LJ. Cognitive Impairment After COVID-19-A Review on Objective Test Data. Front Neurol. 2021;12:699582. Epub 2021/08/17. doi: 10.3389/fneur.2021.699582. PubMed PMID: 34393978.

5. Beaud V, Crottaz-Herbette S, Dunet V, Vaucher J, Bernard-Valnet R, Du Pasquier R, et al. Pattern of cognitive deficits in severe COVID-19. J Neurol Neurosurg Psychiatry. 2021;92(5):567–8. Epub 2020/11/22. doi: 10.1136/jnnp-2020-325173. PubMed PMID: 33219042.

6. Tavares-Junior JWL, de Souza ACC, Borges JWP, Oliveira DN, Siqueira-Neto JI, Sobreira-Neto MA, et al. COVID-19 associated cognitive impairment: A systematic review. Cortex. 2022;152:77–97. Epub 2022/05/11. doi: 10.1016/j.cortex.2022.04.006. PubMed PMID: 35537236.

7. Davis HE, Assaf GS, McCorkell L, Wei H, Low RJ, Re’em Y, et al. Characterizing long COVID in an international cohort: 7 months of symptoms and their impact. EClinicalMedicine. 2021;38:101019. Epub 2021/07/27. doi: 10.1016/j.eclinm.2021.101019. PubMed PMID: 34308300.

8. Rogers JP, Chesney E, Oliver D, Pollak TA, McGuire P, Fusar-Poli P, et al. Psychiatric and neuropsychiatric presentations associated with severe coronavirus infections: a systematic review and meta-analysis with comparison to the COVID-19 pandemic. Lancet Psychiatry. 2020;7(7):611–27. Epub 2020/05/22. doi: 10.1016/S2215-0366(20)30203-0. PubMed PMID: 32437679.

9. D’Esposito M, Postle BR. The Cognitive Neuroscience of Working Memory. Annual Review of Psychology. 2015;66(1):115–42. doi: 10.1146/annurev-psych-010814-015031. PubMed PMID: 25251486.

10. Baddeley AD, Hitch GJ, Allen RJ. From short-term store to multicomponent working memory: The role of the modal model. Mem Cognit. 2019;47(4):575–88. Epub 2018/11/28. doi: 10.3758/s13421-018-0878-5. PubMed PMID: 30478520.

11. Repovs G, Baddeley A. The multi-component model of working memory: explorations in experimental cognitive psychology. Neuroscience. 2006;139(1):5–21. Epub 2006/03/07. doi: 10.1016/j.neuroscience.2005.12.061. PubMed PMID: 16517088.

12. Luck SJ, Vogel EK. Visual working memory capacity: from psychophysics and neurobiology to individual differences. Trends Cogn Sci. 2013;17(8):391–400. Epub 2013/07/16. doi: 10.1016/j.tics.2013.06.006. PubMed PMID: 23850263.

13. Brockmole JR, Logie RH. Age-related change in visual working memory: a study of 55,753 participants aged 8-75. Front Psychol. 2013;4:12. Epub 2013/02/02. doi: 10.3389/fpsyg.2013.00012. PubMed PMID: 23372556.

14. Park DC, Lautenschlager G, Hedden T, Davidson NS, Smith AD, Smith PK. Models of visuospatial and verbal memory across the adult life span. Psychol Aging. 2002;17(2):299–320. Epub 2002/06/14. PubMed PMID: 12061414.

15. Alemanno F, Houdayer E, Parma A, Spina A, Del Forno A, Scatolini A, et al. COVID-19 cognitive deficits after respiratory assistance in the subacute phase: A COVID-rehabilitation unit experience. PLoS One. 2021;16(2):e0246590. Epub 2021/02/09. doi: 10.1371/journal.pone.0246590. PubMed PMID: 33556127.

16. Woo MS, Malsy J, Pottgen J, Seddiq Zai S, Ufer F, Hadjilaou A, et al. Frequent neurocognitive deficits after recovery from mild COVID-19. Brain Commun. 2020;2(2):fcaa205. Epub 2020/12/31. doi: 10.1093/braincomms/fcaa205. PubMed PMID: 33376990.

17. Beresford T, Ronan PJ, Hipp D. A 5-Minute Cognitive Assessment for Safe Remote Use in Patients With COVID-19: Clinical Case Series. JMIR Form Res. 2021;5(6):e26417. Epub 2021/05/20. doi: 10.2196/26417. PubMed PMID: 34010137.

18. Jaywant A, Vanderlind WM, Alexopoulos GS, Fridman CB, Perlis RH, Gunning FM. Frequency and profile of objective cognitive deficits in hospitalized patients recovering from COVID-19. Neuropsychopharmacology. 2021;46(13):2235–40. Epub 2021/02/17. doi: 10.1038/s41386-021-00978-8. PubMed PMID: 33589778.

19. Miskowiak KW, Johnsen S, Sattler SM, Nielsen S, Kunalan K, Rungby J, et al. Cognitive impairments four months after COVID-19 hospital discharge: Pattern, severity and association with illness variables. Eur Neuropsychopharmacol. 2021;46:39–48. Epub 2021/04/07. doi: 10.1016/j.euroneuro.2021.03.019. PubMed PMID: 33823427.

20. Hampshire A, Trender W, Chamberlain SR, Jolly AE, Grant JE, Patrick F, et al. Cognitive deficits in people who have recovered from COVID-19. EClinicalMedicine. 2021;39:101044. Epub 2021/07/29. doi: 10.1016/j.eclinm.2021.101044. PubMed PMID: 34316551.

21. Zhao S, Shibata K, Hellyer PJ, Trender W, Manohar S, Hampshire A, et al. Rapid vigilance and episodic memory decrements in COVID-19 survivors. Brain Commun. 2022;4(1):fcab295. Epub 2022/02/08. doi: 10.1093/braincomms/fcab295. PubMed PMID: 35128398.

22. World Medical A. World Medical Association Declaration of Helsinki: ethical principles for medical research involving human subjects. JAMA. 2013;310(20):2191–4. Epub 2013/10/22. doi: 10.1001/jama.2013.281053. PubMed PMID: 24141714.

23. Schroeder RW, Twumasi-Ankrah P, Baade LE, Marshall PS. Reliable Digit Span: a systematic review and cross-validation study. Assessment. 2012;19(1):21–30. Epub 2011/12/14. doi: 10.1177/1073191111428764. PubMed PMID: 22156721.

24. Tanabe A, Osaka N. Picture span test: measuring visual working memory capacity involved in remembering and comprehension. Behav Res Methods. 2009;41(2):309–17. Epub 2009/04/14. doi: 10.3758/BRM.41.2.309. PubMed PMID: 19363171.

25. Field A. Discovering Statistics Using SPSS. 3rd Edition ed: Sage Publications Ltd., London; 2009.

26. Graham EL, Clark JR, Orban ZS, Lim PH, Szymanski AL, Taylor C, et al. Persistent neurologic symptoms and cognitive dysfunction in non-hospitalized Covid-19 “long haulers”. Ann Clin Transl Neurol. 2021;8(5):1073–85. Epub 2021/03/24. doi: 10.1002/acn3.51350. PubMed PMID: 33755344.

27. Mendez R, Balanza-Martinez V, Luperdi SC, Estrada I, Latorre A, Gonzalez-Jimenez P, et al. Shortterm neuropsychiatric outcomes and quality of life in COVID-19 survivors. J Intern Med. 2021;290(3):621–31. Epub 2021/02/04. doi: 10.1111/joim.13262. PubMed PMID: 33533521.

28. Taquet M, Dercon Q, Luciano S, Geddes JR, Husain M, Harrison PJ. Incidence, co-occurrence, and evolution of long-COVID features: A 6-month retrospective cohort study of 273,618 survivors of COVID-19. PLoS Med. 2021;18(9):e1003773. Epub 2021/09/29. doi: 10.1371/journal.pmed.1003773. PubMed PMID: 34582441.

29. Aghagoli G, Gallo Marin B, Katchur NJ, Chaves-Sell F, Asaad WF, Murphy SA. Neurological Involvement in COVID-19 and Potential Mechanisms: A Review. Neurocrit Care. 2021;34(3):1062–71. Epub 2020/07/15. doi: 10.1007/s12028-020-01049-4. PubMed PMID: 32661794.

30. Doyle MF. Central nervous system outcomes of COVID-19. Transl Res. 2021. Epub 2021/10/04. doi: 10.1016/j.trsl.2021.09.002. PubMed PMID: 34601116.

31. Douaud G, Lee S, Alfaro-Almagro F, Arthofer C, Wang C, McCarthy P, et al. SARS-CoV-2 is associated with changes in brain structure in UK Biobank. Nature. 2022. Epub 2022/03/08. doi: 10.1038/s41586-022-04569-5. PubMed PMID: 35255491.

